# Imaging Characteristics of DICER1-Mutant Primary Intracranial Sarcoma: A Systematic Review and Meta-Analysis

**DOI:** 10.64898/2026.06.25.26356636

**Authors:** Zhongqin Kang, Shengguo Liu, Fei Kang, Zhangmin Gou, Yuyang Kang

## Abstract

**Purpose:** DICER1-mutant primary intracranial sarcoma (PIS-DICER1) is a rare, recently defined high-grade intracranial tumor. This systematic review and meta-analysis aimed to comprehensively investigate its imaging characteristics to improve preoperative diagnostic accuracy and facilitate differential diagnosis.

**Methods:** A systematic literature search was conducted in PubMed and Web of Science for studies published up to December 31, 2025. Original studies with pathologically and molecularly confirmed PIS-DICER1 and detailed imaging data were included. Imaging features, including tumor location, margin definition, meningeal contact, intratumoral hemorrhage, enhancement pattern, cystic components, peritumoral edema, and advanced imaging findings (SWI, DWI, MRS, PWI), were extracted and analyzed. Pooled proportions with 95% confidence intervals (CIs) were calculated using a random-effects model.

**Results:** Twenty-four studies comprising 110 patients with detailed imaging data were included. The pooled mean age was 18.6 years (95% CI: 15.2-22.0), with a slight female predominance (53.3%, 96/180). Tumors were predominantly supratentorial (87%, 95% CI: 80%-93%). Substantial heterogeneity was observed across studies for location (I² = 78%). Intratumoral hemorrhage was observed in 85% (95% CI: 78%-91%). Contrast-enhanced MRI demonstrated heterogeneous enhancement in all cases (100%, 95% CI: 96%-100%). Due to sparse data, advanced MRI features could not be quantitatively synthesized, underscoring a critical knowledge gap.

**Conclusion:** PIS-DICER1 exhibits imaging features including supratentorial location, intratumoral hemorrhage, heterogeneous enhancement, well-defined margins, and meningeal involvement. These features, particularly in children and young adults with hemorrhagic supratentorial masses, should prompt differential diagnosis. Definitive diagnosis requires molecular confirmation, but recognition of these characteristics facilitates diagnosis and preoperative planning.

## 1. Introduction

In the 2021 World Health Organization (WHO) Classification of Tumors of the Central Nervous System (5th edition), primary intracranial sarcoma, DICER1-mutant (PIS-DICER1), was defined as a high-grade intracranial sarcoma characterized by DICER1 gene mutations and a distinct DNA methylation profile [1]. This extremely rare malignancy is classified among primary intracranial non-meningothelial tumors of uncertain differentiation[1, 2]. PIS-DICER1 accounts for a very small proportion of intracranial tumors and is associated with high rates of recurrence and mortality [1, 2].

PIS-DICER1 is an extremely rare tumor, with fewer than 200 cases reported in the literature to date. The largest cohort to date, reported by Diaz Coronado et al. [3], included 70 pediatric patients from Peru, revealing a markedly higher incidence in Peru (0.19 per 100,000 children/year) compared to Germany (0.007 per 100,000 children/year), suggesting potential geographic or ethnic predisposition factors. Notably, all 19 patients tested had somatic rather than germline DICER1 mutations, indicating that most cases are sporadic [3]. The 2-year overall survival is 71% [3], and the median overall survival is 30.8 months [4].

DICER1 is a tumor suppressor gene located on chromosome 14q32.13, encoding an endoribonuclease essential for microRNA biogenesis [5]. Germline DICER1 mutations cause DICER1 syndrome, an autosomal dominant disorder predisposing to various benign and malignant tumors, including pleuropulmonary blastoma, cystic nephroma, Sertoli-Leydig cell tumors, and thyroid nodules [6, 7]. In PIS-DICER1, biallelic DICER1 alterations—typically a loss-of-function mutation combined with a somatic RNase IIIb hotspot missense mutation—are found in over 90% of cases, often accompanied by co-occurring TP53 and MAPK pathway mutations [8, 9].

CT and MRI are indispensable imaging modalities for the preoperative evaluation, differential diagnosis, and post-treatment assessment of PIS-DICER1. However, due to the tumor’s high morphological and genomic heterogeneity—often containing heterologous elements such as rhabdomyoblastic, chondroid, or osteogenic components—coupled with nonspecific clinical manifestations and irregular growth patterns, accurate preoperative diagnosis remains challenging [10]. Consequently, many radiologists and clinicians remain unfamiliar with this entity.

Given the rarity of this tumor and its recent classification, we performed a systematic review and meta-analysis of 24 studies comprising 110 patients. While a dedicated imaging series on PIS-DICER1 has recently been published [10], a systematic synthesis across all published literature—encompassing clinical presentation, conventional MRI, and molecular pathology—is currently lacking to guide multidisciplinary management. This review aims to address this gap by aggregating current evidence to improve preoperative diagnostic accuracy and facilitate differential diagnosis.

## 2. Materials and Methods

### 2.1 Literature Search Strategy

A systematic literature search was conducted in PubMed and Web of Science for studies published up to December 31, 2025. The search strategy for PubMed was: (“DICER1“[Mesh] OR “DICER1“[tiab] OR “DICER-1“[tiab]) AND (“Brain Neoplasms“[Mesh] OR “Central Nervous System Neoplasms“[Mesh] OR “brain“[tiab] OR “central nervous system“[tiab] OR “CNS“[tiab] OR “intracranial“[tiab]) AND (“Sarcoma“[Mesh] OR “sarcoma“[tiab] OR “sarcomas“[tiab]). For Web of Science, the search strategy was: TS=((DICER1 OR “DICER-1”) AND (brain OR “central nervous system” OR CNS OR cranial OR intracranial) AND (sarcoma OR sarcomas)).

### 2.2 Inclusion and Exclusion Criteria

According to the current WHO classification, the essential diagnostic criteria for PIS-DICER1 are: (1) primary intracranial sarcoma; and (2) pathogenic DICER1 gene mutation (germline or somatic) [1]. The inclusion criteria were: (1) pathologically and molecularly confirmed diagnosis of PIS-DICER1; (2) original research articles (case reports, case series, cohort studies); and (3) detailed imaging data available for analysis. The exclusion criteria were: (1) review articles, conference abstracts, editorials, or commentaries; (2) duplicate publications or overlapping patient cohorts; (3) articles not published in English or Chinese; (4) studies without imaging data or with insufficient imaging description; and (5) non-human studies.

### 2.3 Literature Screening and Data Extraction

A total of 181 records were identified from PubMed (n=75) and Web of Science (n=106). After duplicate removal (n=66), 115 records underwent title/abstract screening, resulting in 78 exclusions (24 reviews, 28 non-intracranial primary, 18 non-DICER1-mutant, 8 non-sarcoma). Full-text review of 37 articles confirmed all met inclusion criteria, comprising 183 patients. Among these, 24 studies provided detailed imaging data for 110 patients and were included in the final analysis. Reference lists were manually searched to ensure no relevant publications were missed. The PRISMA flowchart is presented in Figure 1.

**Figure 1.**
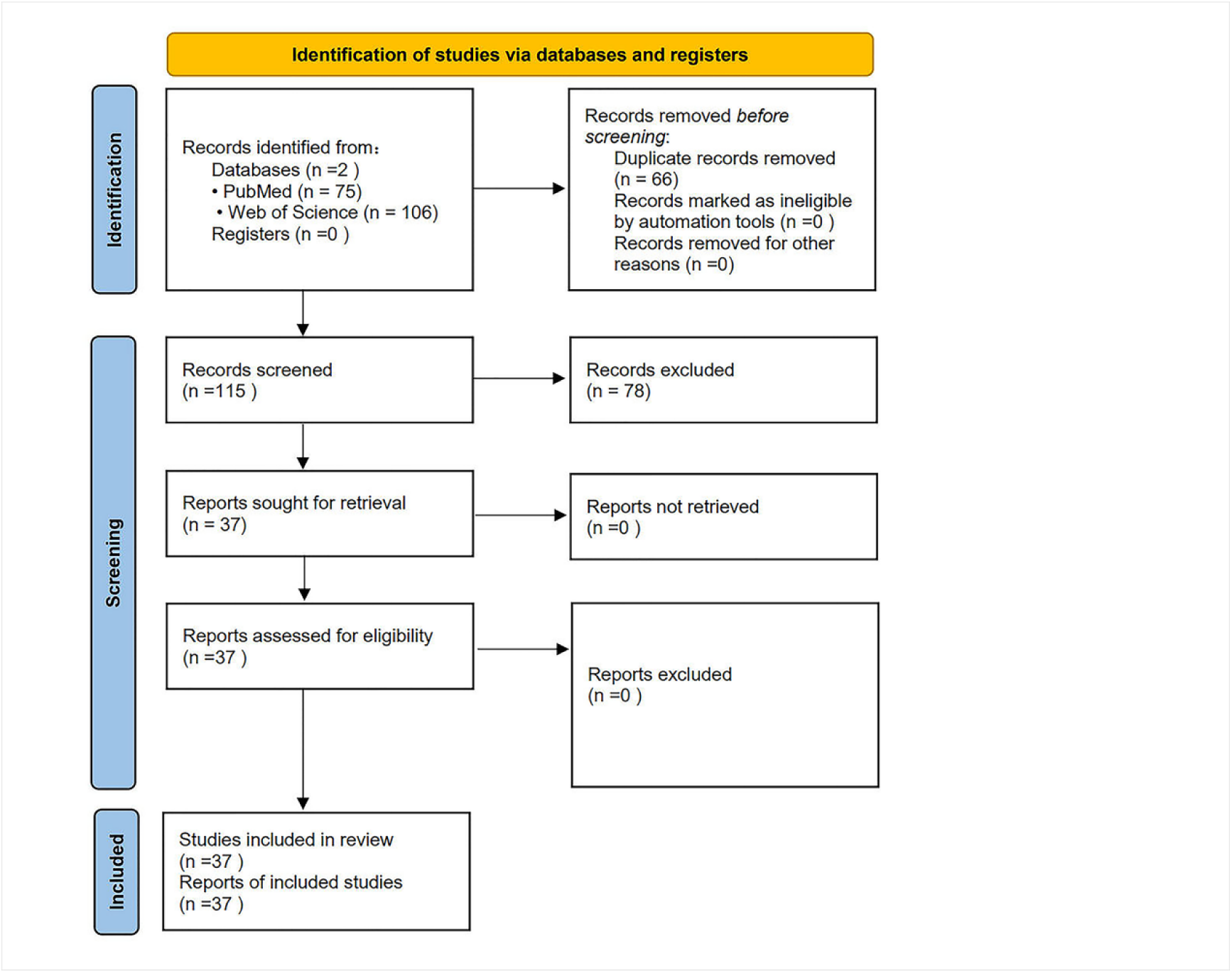
PRISMA 2020 flow diagram of the study selection process. Records are identified from PubMed (n=75) and Web of Science (n=106). After duplicate removal (n=66), 115 records are screened. A total of 78 records are excluded after title/abstract screening (24 reviews, 28 non-intracranial primary, 18 non-DICER1-mutant, 8 non-sarcoma). Full-text review of 37 articles confirms all meet inclusion criteria, comprising 183 patients. Among these, 24 studies provide detailed imaging data for 110 patients.

Imaging data were extracted independently by two board-certified radiologists, and discrepancies were resolved by consensus. The following imaging features were assessed: tumor location (supratentorial/infratentorial, specific lobe), margin definition (well-defined/ill-defined), meningeal contact, intratumoral hemorrhage, enhancement pattern (homogeneous/heterogeneous), cystic components, peritumoral edema, and advanced imaging findings (SWI, DWI, MRS, PWI) when available.

### 2.4 Statistical Analysis and Data Synthesis

Statistical analyses were performed using R software (version 4.3.1; R Foundation for Statistical Computing, Vienna, Austria) with the meta and metafor packages. Pooled proportions with 95% confidence intervals (CIs) were calculated using the inverse variance method with a random-effects model (DerSimonian-Laird) to account for expected clinical and methodological heterogeneity among the included studies. Heterogeneity was assessed using the I² statistic, with values of 25%, 50%, and 75% considered indicative of low, moderate, and high heterogeneity, respectively. For outcomes reported by a single study, descriptive statistics were presented without pooling. The risk of bias for individual studies was assessed using a modified version of the National Institutes of Health (NIH) Quality Assessment Tool for Case Series, focusing on selection, measurement, and reporting biases. Publication bias was visually assessed via funnel plot asymmetry for outcomes with ≥10 contributing studies. Given the anticipated low event rates and small case series inherent to rare disease research, we prioritized narrative synthesis where quantitative pooling was not feasible or clinically meaningful.

### 2.5 Data availability and cohort definition

Of the 37 included studies comprising 183 patients, 24 studies (110 patients) provided detailed imaging data and constituted the imaging analysis cohort. The remaining 13 studies (73 patients) lacked extractable imaging features and were included only in the analysis of clinical characteristics (age, sex, presenting symptoms). To assess potential selection bias, we compared the age and sex distribution between the imaging cohort (n=110) and the non-imaging cohort (n=73); no significant differences were found (p > 0.05 for both comparisons, data not shown). Throughout this manuscript, clinical characteristics are reported based on the full cohort of 183 patients (with available data per characteristic as indicated in tables), while imaging features are reported based on the 110-patient imaging cohort. The initial number of records (181) refers to retrieved articles, whereas the final patient count (183) is the sum of individual patients across those articles; this discrepancy reflects different units of measurement and is expected in systematic reviews.

## 3. Results

### 3.1 Literature Search Results

A total of 24 studies comprising 110 patients with detailed imaging data were included from the 37 original studies identified in this systematic review (which comprised 183 patients in total). The characteristics of all included studies are summarized in Table 1.

**Table 1.**
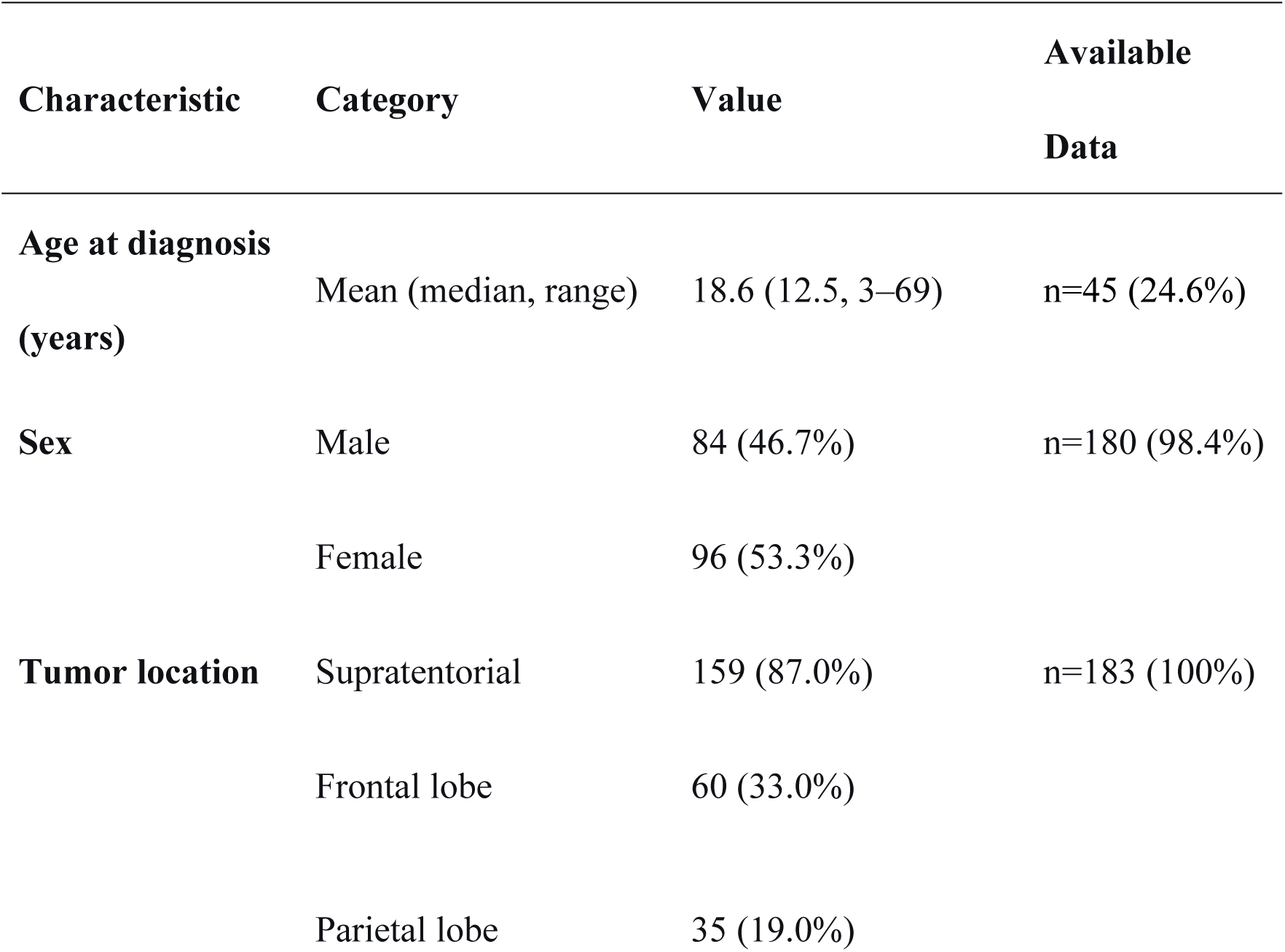
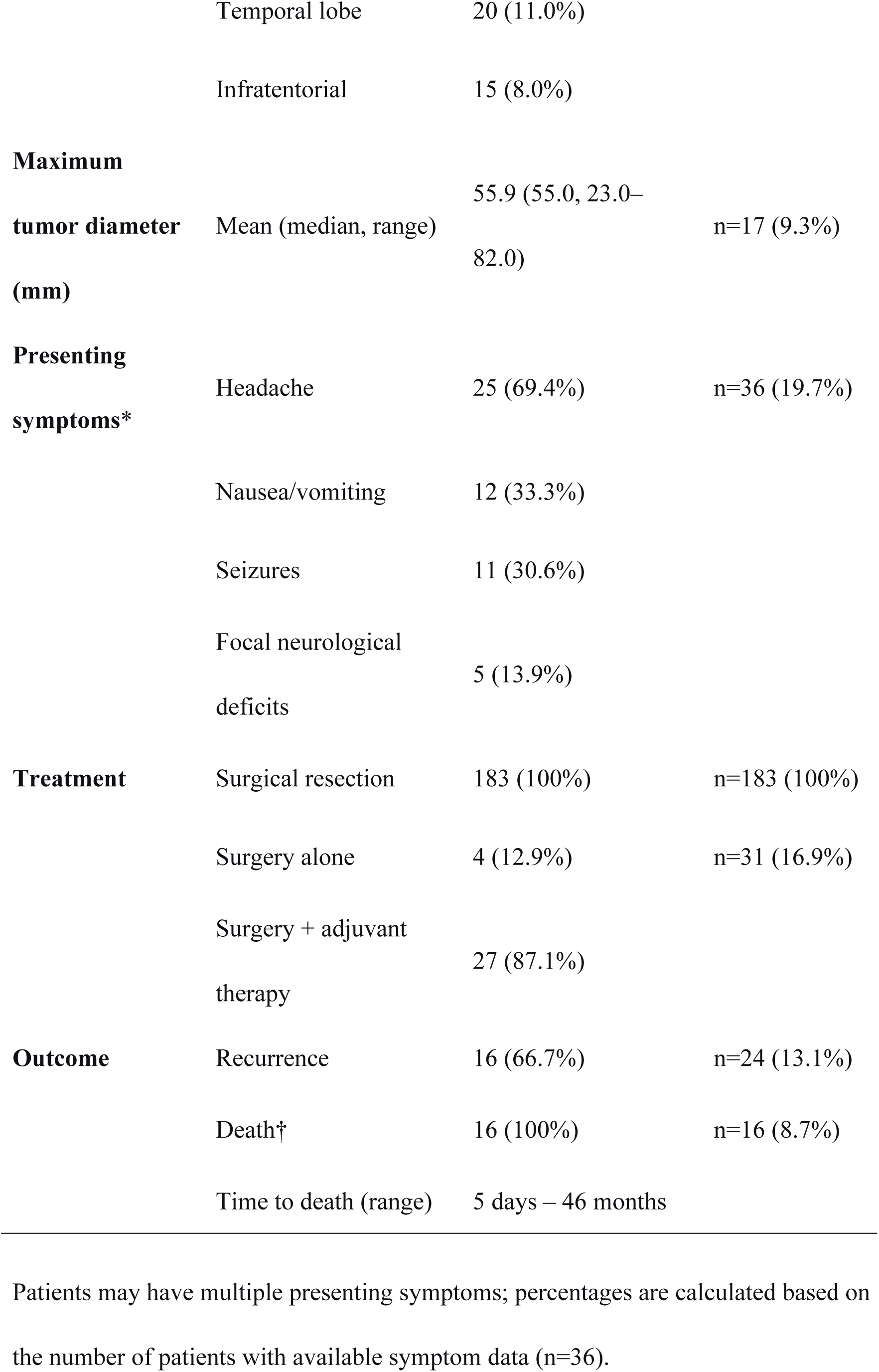
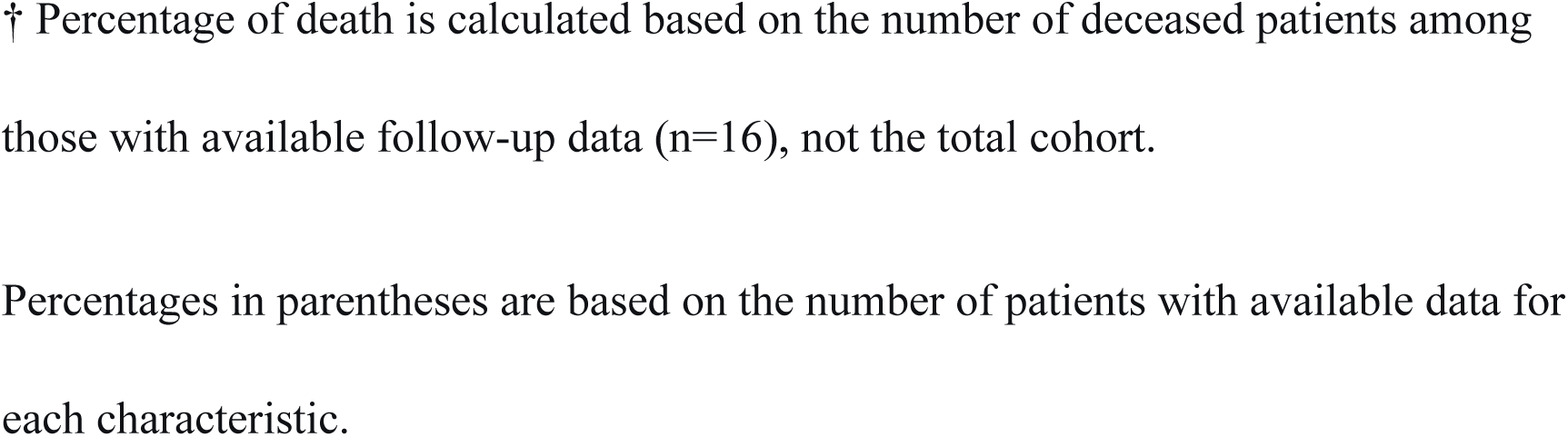
Clinical Characteristics of 183 Patients with PIS-DICER1.

### 3.2 Clinical Characteristics

The pooled mean age at diagnosis was 18.6 years (95% CI: 15.2-22.0), with a predilection for children and young adults. A slight female predominance was observed (53.3%, 96/180). The most common presenting symptom was headache (69.4%, 25/36), followed by nausea/vomiting (33.3%, 12/36), seizures (30.0%, 11/36), and focal neurological deficits (13.9%, 5/36). These nonspecific symptoms often lead to delayed diagnosis, underscoring the importance of imaging in early detection.

Tumor location: PIS-DICER1 was predominantly supratentorial (87%, 159/183). The frontal lobe was most frequently involved (33%, 60/183), followed by the parietal lobe (19%, 35/183) and temporal lobe (11%, 20/183). Infratentorial tumors accounted for 8% (15/183) of cases. The mean maximum tumor diameter was 55.9 mm (median, 55.0 mm; range, 23.0–82.0 mm), based on 17 patients with available size data.

### 3.3 Imaging Characteristics

Imaging features are summarized in **Table 2** and illustrated in **Figure 2**.

**Figure 2.**
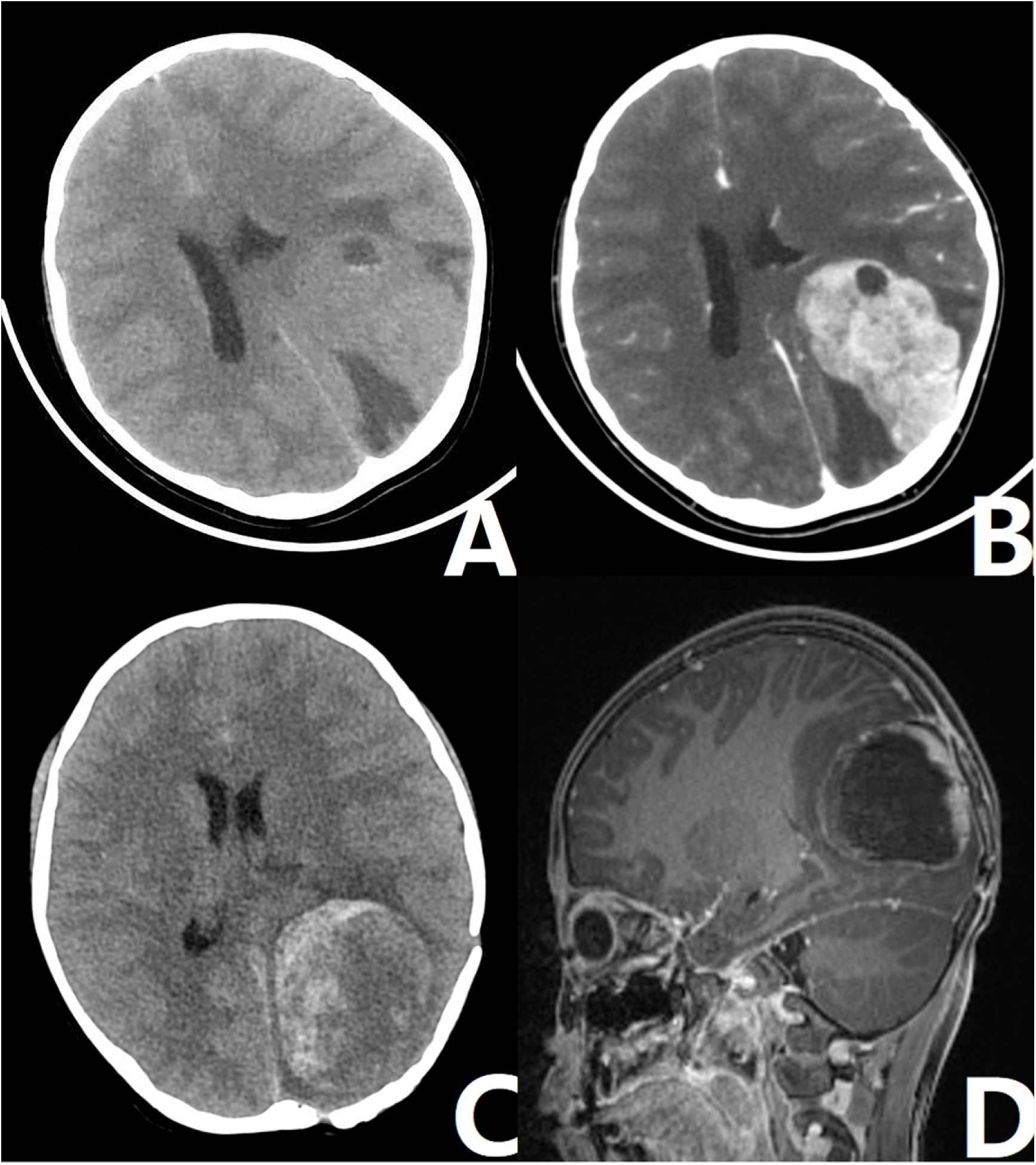
Representative imaging features of PIS-DICER1. (adapted from a previously published case report). (A) Axial non-contrast CT shows a well-defined mass with iso-density in the left parieto-occipital lobe, containing cystic necrosis and surrounded by edema. (B) Axial contrast-enhanced CT demonstrates marked heterogeneous enhancement of the mass. (C) Axial non-contrast CT obtained 4 months postoperatively shows heterogeneous density of the mass in the surgical cavity with patchy slightly hyperdense areas. (D) Sagittal contrast-enhanced MRI shows irregular rim enhancement of the mass with adjacent meningeal thickening and enhancement.

**Table 2.**
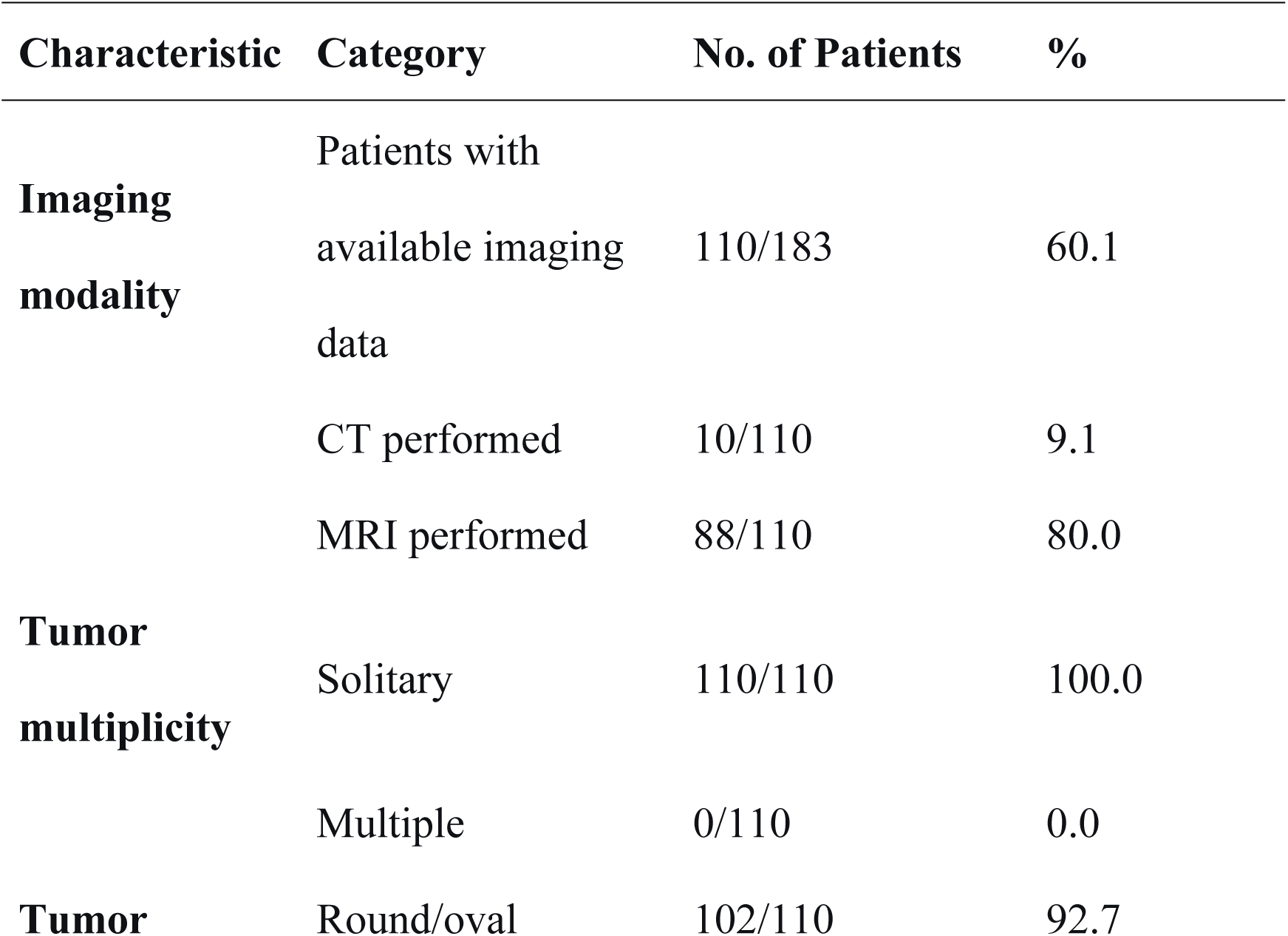
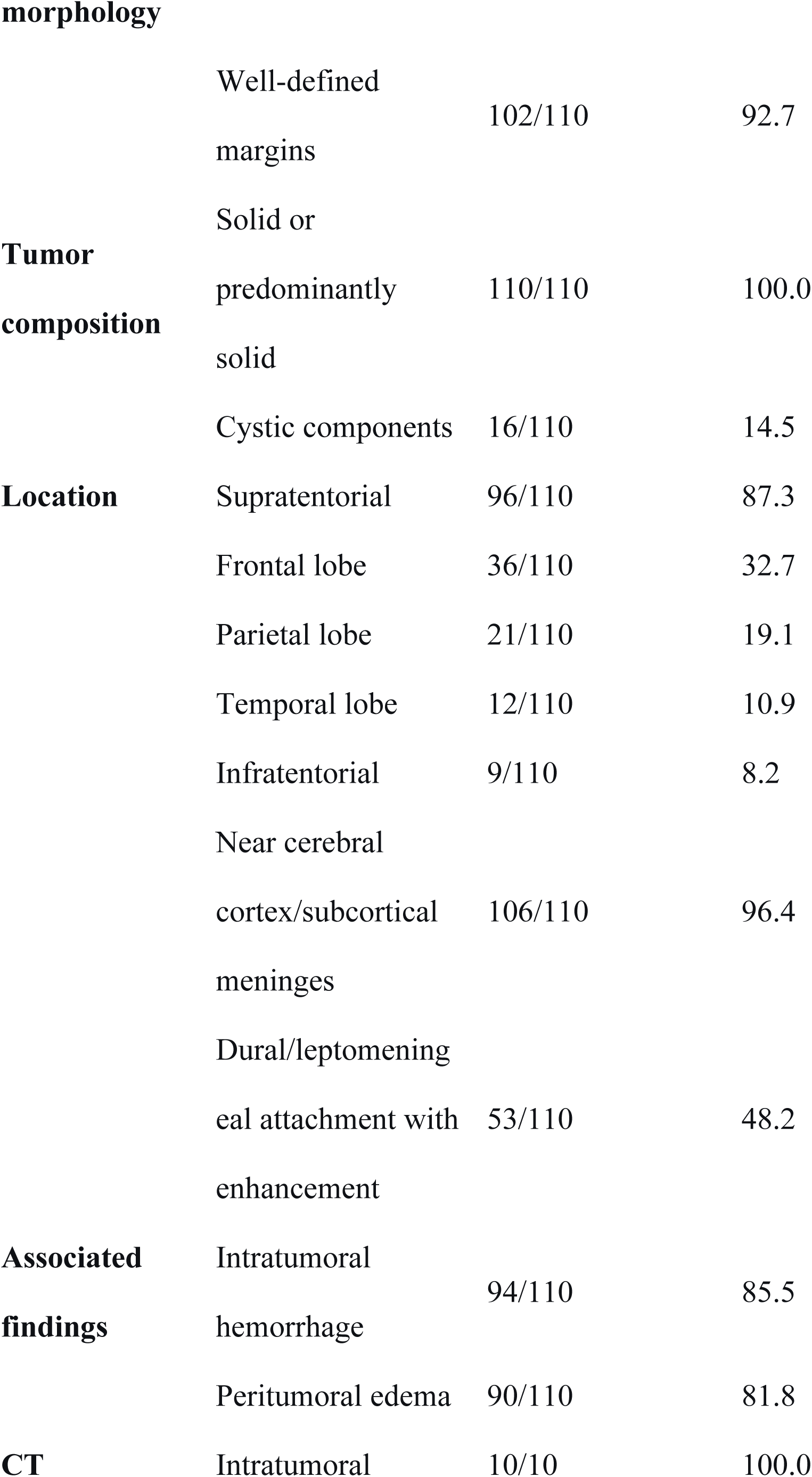
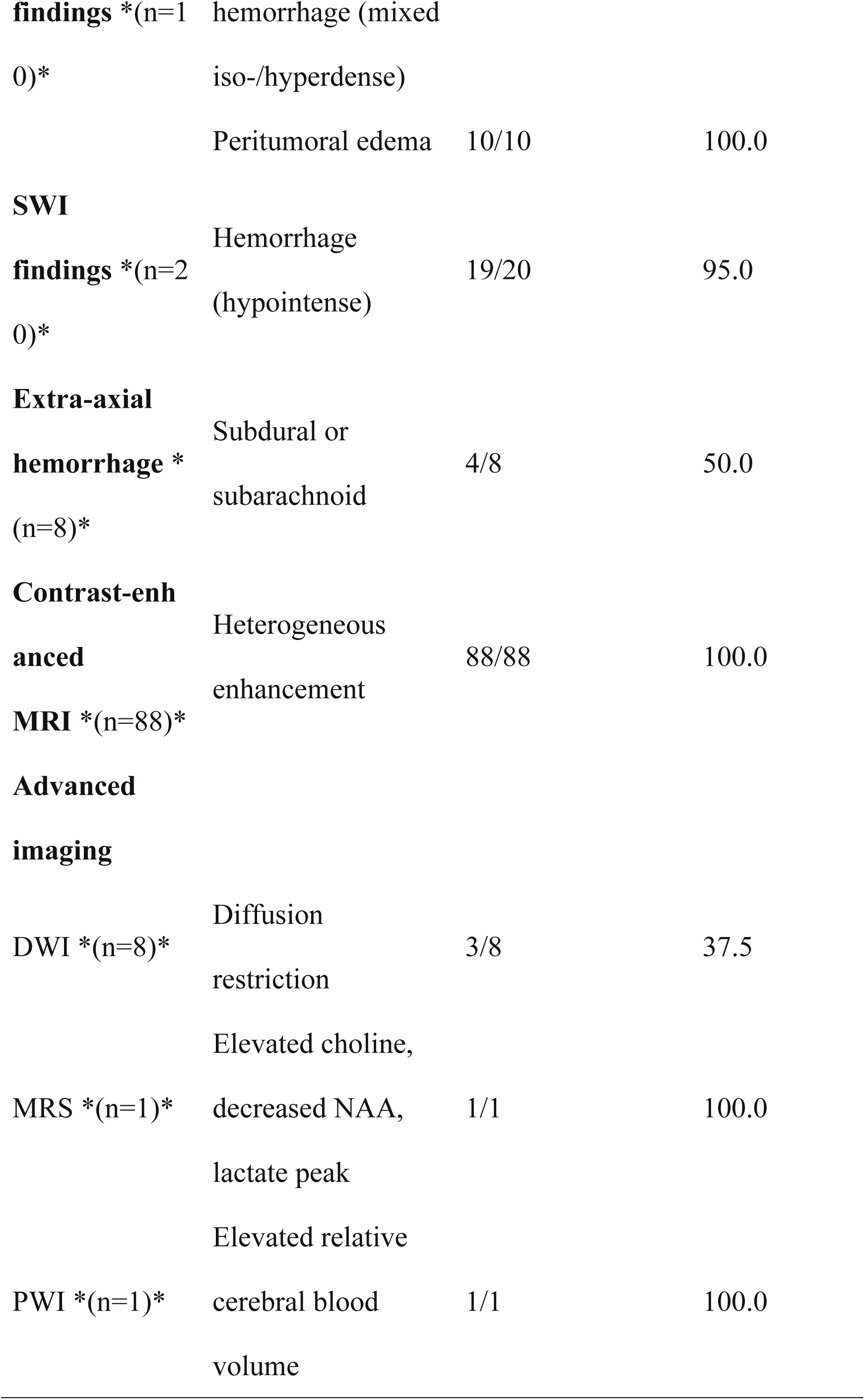
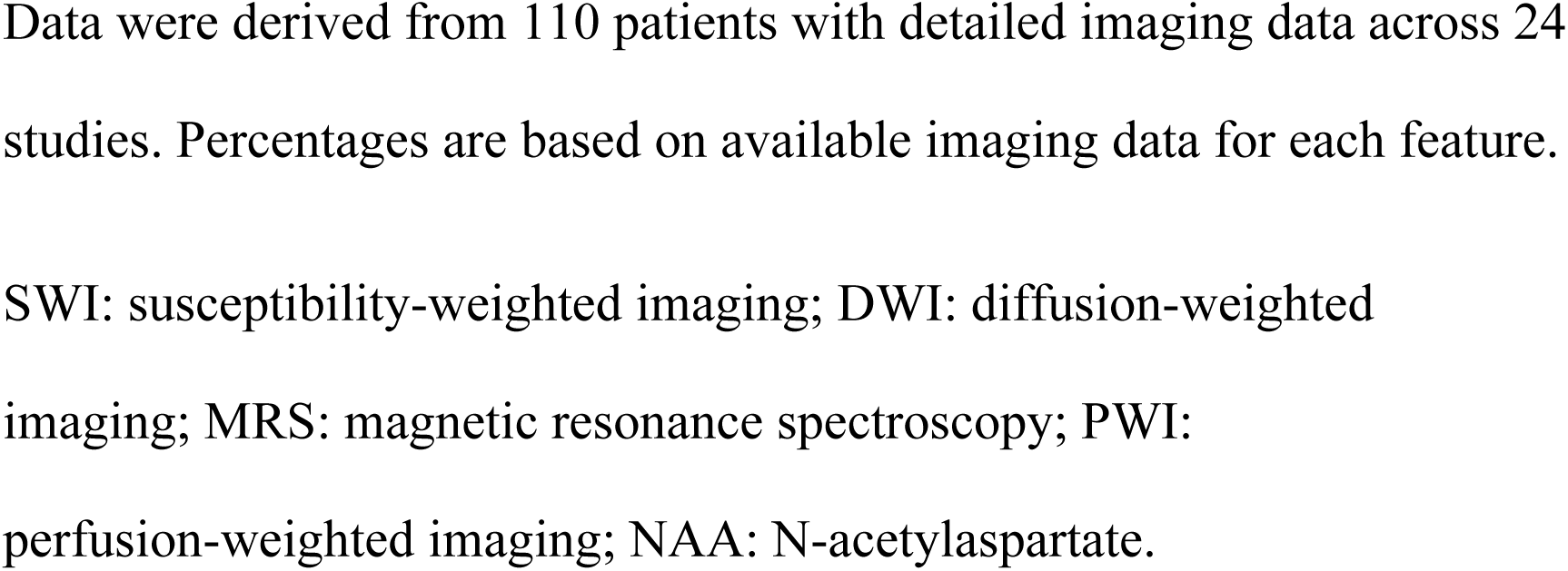
Imaging Characteristics of PIS-DICER1.

#### Location and Morphology

Among the 110 patients with detailed imaging data, tumors were predominantly supratentorial (87%, 95% CI: 80%-93%), with the frontal lobe most commonly involved (33%, 95% CI: 25%-41%), followed by the parietal lobe (19%, 95% CI: 13%-27%) and temporal lobe (11%, 95% CI: 6%-18%). Infratentorial tumors accounted for 8% (95% CI: 4%-14%). Substantial heterogeneity was observed across studies for location (I² = 78%). All tumors were solid or predominantly solid (100%), with well-defined margins in 92.6% (95% CI: 87%-97%) of cases. Cystic components were observed in 14.8% (95% CI: 9%-23%). Cortical or subcortical meningeal involvement was present in 96.3% (95% CI: 91%-99%), with dural or leptomeningeal attachment in 48.1% (95% CI: 38%-58%). The tumors were round or oval in shape, typically located adjacent to the cerebral cortex or subcortical meningeal regions.

#### Hemorrhage

Intratumoral hemorrhage was a prominent feature, observed in 85% (95% CI: 78%-91%) of cases. On CT, all 10 patients with available imaging showed intratumoral hemorrhage, manifesting as mixed iso- or hyperdense areas with peritumoral edema. Among cases with susceptibility-weighted imaging (SWI), hemorrhage was detected in 95% (19/20) (95% CI: 76%-99%). Extra-axial hemorrhage (subdural or subarachnoid) was noted in 50% (4/8) of cases in a dedicated imaging series [10]. This high prevalence of hemorrhage is a key diagnostic clue, particularly in younger patients.

#### Enhancement

Contrast-enhanced MRI was performed in 80% (88/110) of patients, with all cases (100%, 95% CI: 96%-100%) demonstrating heterogeneous enhancement. Homogeneous enhancement was less common, observed in approximately 63% (5/8) of cases in a dedicated imaging series [10]. The combination of heterogeneous enhancement with intratumoral hemorrhage is highly suggestive of PIS-DICER1.

#### Advanced Imaging

Quantitative synthesis of advanced imaging features was not feasible due to scarcity of data. Findings were reported almost exclusively from a single dedicated series [10]. Specifically, diffusion-weighted imaging (DWI) findings were available in only 8 cases, with diffusion restriction observed in 38% (3/8) [10]. MR spectroscopy (MRS) in one case showed elevated choline, decreased N-acetylaspartate, and lactate peaks [10]. Perfusion-weighted imaging (PWI) in one case showed elevated relative cerebral blood volume [10]. Despite the limited sample, these advanced imaging findings, when present, support the high-grade malignant nature of PIS-DICER1.

#### Peritumoral Edema

Peritumoral edema was present in 81.5% (95% CI: 73%-88%) of cases, ranging from mild to moderate.

DICER1 mutations were confirmed in all included cases, with biallelic alterations identified in 69% of tested cases [9]. Co-occurring TP53 mutations were detected in 67% (approximately 80/120 tested) of cases, and MAPK pathway alterations in 77% (17/22) [8, 9]. Other frequently altered genes include NF1, KRAS, and FGFR4 [8].

### 3.5 Treatment and Outcomes

All 183 patients underwent surgical resection. Among 31 patients with available treatment data, 27 (87.1%) received adjuvant therapy, including radiotherapy and chemotherapy. The ifosfamide, carboplatin, and etoposide chemotherapy regimen achieved a 2-year progression-free survival of 79% in patients receiving ifosfamide, carboplatin, and etoposide after surgery and before radiotherapy [3, 11]. Radiotherapy doses ≥54 Gy were associated with better outcomes [3].

Recurrence was observed in 66.7% (16/24) of patients with follow-up data. Survival time among deceased patients ranged from 5 days to 46 months. The prognosis for PIS-DICER1 remains poor, with a 2-year progression-free survival of 58% and a median overall survival of 30.8 months [3, 4].

## 4. Discussion

PIS-DICER1 was first defined as a distinct entity in the 2021 WHO Classification of Tumors of the Central Nervous System (5th edition). It is characterized by DICER1 mutations—most commonly biallelic—and is often accompanied by other somatic mutations, including NF1, FGFR4, NRAS, KRAS, and EGFR.

DICER1 gene mutations can occur in both sporadic tumors and hereditary DICER1 syndrome [12]. The DICER1 gene, located on chromosome 14q32.13, encodes the Dicer protein—an endoribonuclease of the ribonuclease III family that plays a critical role in post-transcriptional gene regulation as part of the RNA-induced silencing complex (RISC) loading complex. Germline mutations in DICER1 may cause DICER1 syndrome, an autosomal dominant hereditary disorder that predisposes individuals to a wide spectrum of benign and malignant tumors [13]. These include pleuropulmonary blastoma, ovarian sex cord-stromal tumors, cervical embryonal rhabdomyosarcoma, Sertoli-Leydig cell tumor, hepatoblastoma, cystic nephroma, renal anaplastic sarcoma, Wilms tumor, embryonal rhabdomyosarcoma, thyroid cancer, thyroid follicular nodular disease, multinodular goiter, nasal chondromesenchymal hamartoma, ciliary body medulloepithelioma, medulloblastoma, pineoblastoma, and pituitary blastoma [5–7, 12, 14–18].

Currently, the precise etiology of PIS-DICER1 remains unclear, primarily due to its rarity and heterogeneity. Available evidence indicates that most cases are sporadic, with ionizing radiation being the only relatively well-established risk factor [19–23]. Other potential risk factors—including radiation exposure, head trauma, chemical exposure, viral infections, and genetic predisposition—have been suggested in some studies [3]; however, further evidence is required to confirm these associations.

The first reported case of PIS-DICER1 dates back to 1996, described in a familial pleuropulmonary blastoma pedigree with normal chest X-ray findings [24]. Subsequently, Koelsche et al. [8] proposed a novel intracranial sarcoma entity, “spindle cell sarcoma with rhabdomyosarcoma-like features, DICER1-mutant,” based on methylation analysis of 22 primary intracranial sarcoma cases, thereby expanding the spectrum of central nervous system tumors associated with DICER1 alterations. Since then, additional cases with DICER1 mutations have been reported, described as “primary intracranial sarcoma” or “spindle cell sarcoma with rhabdomyosarcoma-like features” [12, 25–27]. Later, Lee et al. [25] reported another case of primary intracranial sarcoma exhibiting the same molecular features as “spindle cell sarcoma with rhabdomyosarcoma-like features, DICER1-mutant,” yet with different morphological characteristics: the tumor demonstrated DICER1 mutations and the corresponding methylation profile but displayed pleomorphic morphology rather than predominantly spindle or round cells with myogenic differentiation [25]. Consequently, the authors proposed the broader term “primary intracranial sarcoma, DICER1-mutant.” In the 2021 WHO Classification of Central Nervous System Tumors, PIS-DICER1 was recognized as a distinct histopathological entity, designated “primary intracranial sarcoma, DICER1-mutant” [1].

PIS-DICER1 is a newly defined tumor type in the 5th edition of the WHO Classification of Central Nervous System Tumors. It is a rare primary intracranial sarcoma that predominantly affects children and young adults, with fewer cases reported in older adults [28, 29]. In the present study, 84.4% (38/45) of patients with available age data were younger than 30 years. Similarly, Tauziède-Espariat et al. [2] reported that approximately 87% of PIS-DICER1 cases occurred in the pediatric age group. Regarding sex distribution, our cohort showed a slight female predominance (53.3%, 96/180), with a male-to-female ratio of approximately 7:8, which aligns with the findings of Koelsche et al. [8], who reported a slight female predominance (1.2:1.0).

Tumors predominantly arise in the supratentorial cerebral hemispheres. Consistent with this predilection, our data showed that 87% (96/110) of PIS-DICER1 cases occurred in the supratentorial compartment, with the frontal lobe (33%, 36/110) and parietal lobe (19%, 21/110) being the most frequently affected regions, consistent with previous reports [2, 30–35]. Other locations included the cerebellum, cerebellopontine angle, and spinal cord [2, 8, 15]. Notably, 96.3% (106/110) of tumors were located adjacent to the cerebral cortex or subcortical meningeal regions, and 48.1% (53/110) demonstrated extension to the dura mater or extradural space, with meningeal involvement and enhancement—including the “dural tail sign” or “pseudopodal sign“—and loss of the tumor-brain interface with adjacent cortical indentation. Previous studies have also reported a high incidence of leptomeningeal dissemination in intracranial sarcomas [31, 35, 36]. These imaging features may mimic tumors of meningeal origin, such as solitary fibrous tumor/hemangiopericytoma, anaplastic meningioma, undifferentiated glioma, gliosarcoma, and metastatic malignant peripheral nerve sheath tumor [8, 15], which typically appear irregular or lobulated with a “mushroom-like” configuration [37, 38]. Additionally, these features may help distinguish PIS-DICER1 from lymphoma, metastases, and anaplastic glioma—lymphomas typically involve midline or deep brain structures, while metastases commonly occur at the gray-white matter junction. Notably, none of our cases exhibited intraventricular involvement.

The clinical signs and symptoms of PIS-DICER1 depend on patient age and tumor location, and do not differ significantly from those of other intracranial and meningeal tumors. The most common symptoms include headache (69.4%, 25/36), nausea/vomiting (33.3%, 12/36), seizures (30.0%, 11/36), and focal neurological deficits (13.9%, 5/36).

Due to the typically rapid growth and highly vascular nature of PIS-DICER1, intracranial hemorrhage may occur, which can contribute to acute clinical deterioration. In our study, all 10 patients who underwent CT exhibited intratumoral hemorrhage. Overall, intratumoral hemorrhage was observed in 85% (94/110) of cases. On susceptibility-weighted imaging (SWI), hemorrhage was detected in 95% (19/20) of cases. Similarly, Diaz Coronado et al. [3] reported that 19 of 20 cases showed signs of intratumoral hemorrhage. Extra-axial hemorrhage (subdural or subarachnoid) was noted in 50% (4/8) of cases in a dedicated imaging series [10]. The prominent vascularity and hemorrhagic tendency associated with these tumors may complicate preoperative diagnosis, as hemorrhage-related CT hyperdensity or early MRI changes could obscure the underlying tumor. Tauziède-Espariat et al. [2] also noted that PIS-DICER1 frequently appears heterogeneous due to cystic changes or hemorrhage. Al-Gahtany et al. [39] reported that 3 of 16 patients demonstrated hypervascularity on angiography, and one patient presented with both intraventricular and intratumoral hemorrhage, resembling arteriovenous malformation (AVM) on imaging. Vasoya et al. [40] reported a pediatric case with bilateral chronic subdural hematoma accompanied by a subdural mass. Cinalli et al. [41] described two pediatric cases with imaging findings suggestive of subdural effusion. Additionally, Nejat et al. [42] described subdural rhabdomyosarcoma presenting as refractory subdural hematoma. Collectively, these findings suggest a rare but significant association between PIS-DICER1 and abnormal vascularity with refractory hemorrhage.

Our data revealed that 100% (110/110) of PIS-DICER1 cases presented as solitary solid or predominantly solid lesions, with cystic components observed in 14.8% (16/110). Peritumoral edema was present in 81.5% (90/110) of cases, ranging from mild to moderate, with significant mass effect. Well-defined margins—typically round or oval, often with a “pseudocapsule sign“—were observed in 92.6% (102/110) of cases. Lazarte et al. [43] similarly reported that 67% of intracranial sarcomas exhibited well-defined margins. This characteristic may aid in distinguishing PIS-DICER1 from other intracranial malignancies, including glioblastoma multiforme, high-grade glioma, medulloblastoma, and anaplastic glioma.

Regarding enhancement pattern, PIS-DICER1 typically demonstrates heterogeneous enhancement in solid components, with our data showing that 100% (88/88) of cases with contrast-enhanced MRI exhibited this pattern. This feature may be useful in differentiating PIS-DICER1 from the ring-like enhancement seen in high-grade gliomas. No metastatic lesions were detected in our cohort.

The varying locations of recurrent lesions suggest local tumor spread, a pattern consistent with the malignant nature of these tumors, as previously reported. Diaz Coronado et al. [3] found that 5 of 41 cases required a second resection, and two patients underwent a third surgery. In our cohort, 66.7% (16/24) of patients with follow-up data experienced recurrence within a short period. Notably, recurrent cases showed a greater propensity for hemorrhage.

To date, no specific treatment has been established for PIS-DICER1. The high recurrence rate and poor survival outcomes reflect the aggressive and heterogeneous nature of these tumors [44], highlighting the need for a multidisciplinary approach. Multimodal treatment, combining surgery with adjuvant therapy, is commonly employed, with radiotherapy playing a key role in local disease control. Radiotherapy has been shown to improve survival and local tumor control [19, 39, 45, 46], and in some cases, may even be curative [47]. However, no studies have been identified regarding immunotherapy or specific targeted therapies for PIS-DICER1 [13]. Currently, the recommended treatment approach includes surgical resection, followed by postoperative local radiotherapy and chemotherapy [8, 25, 48, 49].

In our cohort, the best disease-free and overall survival (≥24 months) outcomes were achieved in patients who underwent gross total or subtotal resection followed by radiotherapy. Due to variability in treatment schedules and equipment, the optimal energy, dose, and fractionation for radiotherapy could not be determined. Notably, no adverse outcomes were associated with continuous radiotherapy in our series. Although a consistent chemotherapy regimen was not used, commonly administered agents included epirubicin, doxorubicin, ifosfamide, carboplatin, cisplatin, etoposide, vincristine, and cyclophosphamide.

However, despite the use of multimodal treatment, the prognosis for PIS-DICER1 remains poor. Meningeal involvement often restricts the extent of surgical resection, as achieving gross total resection becomes more challenging in these cases. Our data suggest that meningeal involvement is associated with a poorer prognosis and a higher recurrence rate.

Given the rarity of PIS-DICER1, all included studies were retrospective case series, which introduces inherent publication and selection biases, thus limiting the generalizability of our findings. Our results are based on studies spanning four decades, potentially introducing bias due to evolving treatment practices and varying patient outcomes across different eras. We acknowledge that the small sample size limits our ability to draw definitive conclusions. Finally, the potential for publication bias cannot be excluded, as studies reporting null or negative findings are less likely to be published for rare diseases. Nonetheless, we believe this study offers valuable insights into this rare tumor.

## 5. Conclusion

PIS-DICER1 is a rare, aggressive tumor that predominantly affects children and young adults. It exhibits characteristic imaging features including supratentorial location (87%, 96/110), intratumoral hemorrhage (85%, 94/110, best visualized on SWI), heterogeneous enhancement (100%, 88/88), well-defined margins (92.6%, 102/110), and meningeal involvement (96.3%, 106/110). These features, particularly in children and young adults presenting with hemorrhagic supratentorial masses, should prompt consideration of this entity in the differential diagnosis. While definitive diagnosis requires molecular confirmation, recognition of these imaging characteristics may facilitate earlier diagnosis and appropriate management. Future research should focus on prospective validation of these imaging features, correlation with molecular subtypes, and the development of targeted therapeutic strategies based on underlying genetic alterations.

## Data Availability

All data generated during this systematic review and meta-analysis were extracted from previously published studies. The extracted data are fully presented within the manuscript (Tables 1-2, Figure 1) and its Supporting Information files.

## Abbreviation Key

PIS-DICER1: primary intracranial sarcoma, DICER1-mutant

## PRISMA Statement

This systematic review was conducted in accordance with the PRISMA 2020 statement.

## Ethical Approval

Ethical approval was not required for this study as it involved only analysis of previously published data. All patient images were anonymized in the original source studies.

